# Use of autologous platelet derivatives for secondary alveoloplasty in patients with cleft lip and palate: a protocol for a systematic review and meta-analysis

**DOI:** 10.1101/2021.06.28.21259643

**Authors:** Andrés Campolo, Claudia Heider, Ismael Cañete, María Francisca Verdugo, Rocio Bravo Jeria, Carmen Gloria Morovic, Gabriel Rada

## Abstract

**Objective:** To assess the effectiveness and safety of autologous platelet derivatives, specifically platelet-rich plasma (PRP) or platelet-rich fibrin (PRF), for secondary alveoloplasty in patients with cleft lip and palate.

**Eligibility criteria:** We will include randomized trials evaluating the effect of autologous platelet derivatives on newly bone formed after secondary alveoloplasty in cleft lip and palate patients. Two reviewers will independently screen each study for eligibility, data extraction, and bias assessment using the Cochrane “risk of bias “tool. We will pool the results using meta-analysis and apply the GRADE system to assess the certainty of the evidence for each outcome.

**Data sources:** A comprehensive search will include all relevant randomized controlled trials (RCTs), the ongoing investigation reported in specialty congresses and trials regardless of language or publication status (published, unpublished, in press, and progress). We will conduct searches in the following databases: Cochrane Central Register of Controlled Trials (CENTRAL), PUBMED, Embase, and LILACS. We will screen trial registries and other sources in order to identify articles that might have been missed in the electronic searches.

**Ethics and dissemination:** As researchers will not access information that could identify an individual participant, obtaining ethical approval was waived.

## INTRODUCTION

### Rationale

The alveolar cleft is a common congenital anomaly that affects approximately 75% of cleft lip and palate patients^1^. This condition can affect the developing dentition and contribute to the collapse of the alveolar segments. The untreated alveolar cleft can also develop other dysfunctions, including oronasal fistula, fluid reflux into the nasal cavity, speech disorders, deficiency of the maxilla in the anteroposterior and transverse direction, lack of bone support for the anterior teeth, dental crowding, and facial asymmetry^2^.

The treatment for the closure of the alveolar cleft is secondary alveoloplasty, which consists of a bone graft technique to fill the defect between both maxilla segments. Since the first description of secondary alveolar bone grafting (SABG) by Boyne and Sands in the 1970s, this intervention has become the most acknowledged method for repairing alveolar clefts^3^. Some recent modifications of the technique seek to improve the percentage of new bone formed and bone density^4,5^.

In recent years, regenerative medicine has emerged as an alternative method for treating bone defects, including treating alveolar cleft through cell therapy, autologous platelet derivatives (APD), either as platelet-rich fibrin (PRF) or platelet-rich plasma (PRP), and scaffolds^5^. Among the advantages of APD is that they contain abundant growth factors, such as platelet-derived growth factor (PDGF), vascular endothelial growth factor (VEGF), and transforming growth factor-beta (TGF-b)^6^. Theoretically, these factors can promote bone growth and reduce bone resorption after surgery^6,7^. Nevertheless, these results are still controversial, and there is no conclusive information in this regard^8,9^.

Even though available evidence on autologous platelet derivatives as an adjunct to bone grafting to treat alveolar cleft has increased in the last decade, to our best knowledge, there is no systematic review that has carried out a synthesis and analysis of the available literature about this topic.

### Objective

To evaluate the effectiveness and safety of autologous platelet derivatives (PFR or PRP) as an adjunct with autologous bone graft to repair alveolar clefts compared with the conventional technique of secondary alveolar bone grafting.

## METHODS

### Types of studies

We will only include randomized controlled trials. We will exclude studies evaluating the effects on animal models or in vitro conditions.

### Types of participants

We will include trials assessing participants meeting the following criteria: patients of all ages with alveolar cleft undergoing secondary alveoloplasty, including patients with a unilateral or bilateral cleft.

### Types of interventions

We will include trials evaluating the use of platelet-rich plasma or platelet-rich fibrin, an adjunct with autologous bone graft to repair alveolar clefts. The APD must be mixed with the bone graft and then used to fill the maxillary gap as the final surgery step before wound closure. We will exclude studies using APD with any technique other than those listed above.

The intervention will be compared with the conventional technique of secondary alveolar bone graft using autologous bone.

We will exclude studies whose control group received other types of grafts (allografts, xenografts, scaffolds).

### Types of outcome measures

We will not use outcomes as an exclusion criterion during the selection process. Any article meeting all the criteria except for the outcome criterion will be preliminarily included and evaluated in full text. We will consider the following outcomes as relevant:

#### Primary outcomes

- Functionality: These types of outcomes include but are not limited to, oronasal fistula, fluid reflux into the nasal cavity, speech disorders, deficiency of the maxilla in the anteroposterior and transverse direction, lack of bone support for the anterior teeth, and dental crowding.
- Quality of life

#### Secondary outcomes

- Postoperative complications such as reoperation, infection, postoperative pain, inflammation, dehiscence, periodontal problems among others.

#### Other outcomes

- Bone volume (newly formed bone) using three-dimensional imaging modalities.
- Bone density using three-dimensional imaging modalities.
- Mean bone loss in height or width.

### Search methods for identification of studies

The searches will cover the inception date of each database and the end date will be one week before submission. No study design, publication status, or language restriction will be applied.

### Electronic searches

We will search the Cochrane Central Register of Controlled Trials (CENTRAL), PUBMED, Embase, LILACS, WHO International Clinical Trials Registry Platform (WHO-ICTRP) (www.who.int/ictrp/), ClinicalTrials.gov (clinicaltrials.gov/) and Google Scholar databases without language or date restrictions. Grey literature searches will be conducted to identify studies not indexed in the databases listed above in OpenGrey (www.opengrey.eu/). The search strategy is presented in supplementary file 1.

### Searching other resources

We will review the reference lists of all included studies and relevant systematic reviews for additional potentially eligible primary studies. Additionally, we will contact the authors of eligible studies and researchers with expertise pertinent to the review topic. We will conduct cross-citation searches via Google Scholar and search in not indexed specialized journals in the field of oral and maxillofacial surgery. Abstracts and oral presentations of specialty meetings and congress will be reviewed as well: American Cleft Palate-Craniofacial Association (ACPA) Annual Meeting, American Society of Craniofacial Surgeons (ASCFS) Annual Meeting, European Cleft Palate Craniofacial Association (ECPCA) Congresses, European Association for Cranio Maxillo Facial Surgery (EACMFS) Congresses, Spanish Society of Facial Fissures (SOCEFF) Congresses, Latin American Craniofacial Association (LATICFA) Congresses, European Society of Craniofacial Surgery (ESCFS) Meetings, Mexican Association of Cleft Lip, Palate and Craniofacial Anomalies (AMLPHAC) Congresses, International Confederation of Cleft Lip and Palate and Related Craniofacial Anomalies (ICCPCA) Congresses, International Society of Craniofacial Surgery (ISCFS) Congresses, Brazilian Society of Craniomaxillofacial Surgery (ABCCMF) and Brazilian Society of Cleft Lip and Palate (ABFLP) congresses, International Comprehensive Cleft Care Workshop (CCCW) Conferences.

### Data collection and analysis

#### Selection of studies

Two review authors will independently screen titles and abstracts of all articles yielded by the searches against the inclusion criteria. We will obtain full texts for all articles that seem to fulfill inclusion criteria, and review authors will independently assess them to decide on their inclusion. Duplicates will be identified by comparing authors of the reports, trial dates, trial durations, number of participants, details on the interventions, location, and setting of the reported trials and will be removed using Rayyan software^10^ Disagreements will be solved through discussion. In case of no agreements, a third author will decide. Articles retrieved from the screening and included in the review will be recorded in the RevMan software^11^. After full-text revision, excluded trials and their primary reason for the decision will be listed.

We will document the selection process in sufficient detail to complete a PRISMA flow chart^12^ and a table of ‘Characteristics of excluded studies ‘ as recommended in Section 11.2.1 of the Cochrane Handbook for Systematic Reviews of Intervention^13^.

### Data extraction and management

Two review authors will independently extract data using a standardized form and check for agreement before data entry into Review Manager 5^11^. For each study with more than one control or comparison group for the intervention, other than “intervention “, we will extract only the results of the control group “with no intervention. “We will not double count data within a meta-analysis.

If different types of PRP or PRF are to be found, they will be considered into one single intervention. The following details will be recorded for each trial in the data extraction form: study design; country of origin; recruitment period; funding source; inclusion criteria; exclusion criteria; the number of patients randomized number of patients evaluated;age of participants, interventions (e.g., conventional treatments, preventive measures, no intervention); platelet derivative preparation, duration of interventions; primary and secondary outcomes; time points were outcomes were measured; method of sample size calculation; duration of follow-up; whether groups were comparable at baseline in relation to diagnosis; and any co-interventions.

### Assessment of risk of bias in included studies

Included studies will be assessed independently by two review authors using the revised Cochrane risk of bias tool (RoB 2)^14^. Any disagreements will be resolved by discussion.

We will evaluate each trial for the following domains:

1. Bias arising from the randomization process.
2. Bias due to deviations from intended interventions.
3. Bias due to missing outcome data.
4. Bias in measurement of the outcome.
5. Bias in selection of the reported result.

Each domain will be judged in terms of high, unclear, or low risk of bias, as described in Chapter 8 of the Cochrane Handbook^12^. These domains will inform an overall risk of bias, defined as follows:

- Low risk of bias: The study is judged to be at low risk of bias for all domains for this result.
- Some concerns: The study is judged to be at some concerns in at least one domain for this result.
- High risk of bias: The study is judged to be at high risk of bias in at least one domain for this result, or the study is judged to have some concerns for multiple domains in a way that substantially lowers confidence in the result

The assessments for each included study will be reported using RevMan 5^10^.

### Measures of treatment effect

For dichotomous outcomes, risk ratios (RR) along with 95% confidence intervals (CI) will be estimated. For continuous outcomes, weighted mean differences (WMD) and standard deviation along with 95% CI will be pooled when trials use the same scale. For continuous outcomes using different scales, the standardized mean difference (SMD) with 95% CI will be used.

### Dealing with missing data

For missing data (for example, publication bias, an outcome not measured, lack of intention-to-treat (ITT) analysis, attrition from the study), the following strategies would be adopted:

- Whenever possible, contact the original investigators to request missing data.
- Make explicit the assumptions of any methods used to cope with missing data: for example, that the data are assumed missing at random or that missing values were assumed to have a particular value, such as a poor outcome.
- Perform sensitivity analyses to assess how sensitive results are to reasonable changes in the assumptions that are made.
- Address the potential impact of missing data on the findings of the review in the ‘Discussion ‘ section.

### Assessment of heterogeneity

Clinical heterogeneity will be assessed by two authors, and meta-analysis will be only conducted where both agree that study participants, interventions, and outcomes are sufficiently similar. We will assess statistical heterogeneity by visually inspecting the forest plots, by considering the Chi^2^ test (with significance level set at P value < 0.10), and using the I^2^ statistic. Where statistical heterogeneity is moderate, substantial, or high (I^2^ > 75%) or where there is clinical heterogeneity, we will investigate possible causes by exploring th*e* impact of participants ‘ characteristics or other variables.

### Assessment of reporting biases

We will assess reporting bias by comparing outcomes reported in the published report against the study protocol whenever this could be obtained. If we could not obtain the protocol, we will compare outcomes listed in the methods section with those whose results were reported. If information is insufficient to judge the risk of bias, we will note this meta-analysis as having some concerns. If any meta-analysis includes a sufficient number of trials (more than 10), we will assess publication bias according to the recommendations on testing for funnel plot asymmetry, as described in Section 10.4 of the Cochrane Handbook for Systematic Reviews of Interventions^13^. If asymmetry is identified, we will examine possible causes or assess the asymmetry by using a table to list the outcomes reported by each study included in the review to identify whether any studies did not report outcomes that had been reported by most studies.

### Data synthesis

The results of clinically homogeneous studies will be meta-analyzed using the statistical package Review Manager 5, provided by Cochrane^11^. We will combine risk ratios for dichotomous data and mean differences for continuous data using the inverse variance method with the random-effects model. For any outcomes for which the included studies are not sufficiently homogeneous or where insufficient data for meta-analysis are found, a narrative synthesis will be presented.

### Subgroup analysis and investigation of heterogeneity

The following sub-groups will be investigated, if possible:

– Type of APD used (PRF -PRP)
– Type of cleft lip and palate (unilateral, bilateral)

In case we identify significant differences between subgroups (test for interaction <0.05) we will report the results of individual subgroups separately.

### Sensitivity analysis

We will perform sensitivity analysis excluding studies with a high risk of bias. In cases where the primary analysis effect estimates and the sensitivity analysis effect estimates significantly differ, we will either present the low risk of bias -adjusted sensitivity analysis estimates -or present the primary analysis estimates but downgrading the certainty of the evidence because of the risk of bias.

### Presenting results and ‘Summary of findings ‘ tables

We will summarize the findings of the main intervention comparison by undertaking the following procedure to create a “Summary of Findings “(SoF) table:

- Assessing the certainty of the evidence for each outcome using the GRADE approach
- Summarizing the findings for each outcome (quantitatively, where possible).
- Completing the SoF table.
- Preparing bullet points that summarize the information in the SoF table in plain language.

Review authors will independently assess the certainty of the evidence (high, moderate, low, and very low) using the five GRADE considerations (study limitations, consistency of effect, imprecision, indirectness, and publication bias). We will use methods and recommendations described in Section 8.5 and Chapter 12 of the Cochrane Handbook^13^ and GRADEpro software^15^ to build SoF tables. We will resolve disagreements on certainty ratings by discussion, provide justification for decisions regarding the ratings using footnotes in the table, and make comments to aid readers ‘ understanding of the review where necessary. We will use plain language statements to report these findings.

## Data Availability

This is a systematic review, and all the data included in this study is available from Cochrane Central Register of Controlled Trials (CENTRAL), PUBMED, Embase, LILACS, WHO International Clinical Trials Registry Platform (WHO-ICTRP) (www.who.int/ictrp/), ClinicalTrials.gov (clinicaltrials.gov/) and Google Scholar databases.

## Ethics and dissemination

As researchers will not access information that could identify an individual participant, obtaining ethical approval was waived.

## Authors ‘ contributions

A.C., C.H., I.C., M.V., R.B., and C.M. conceived the protocol. A.C. and C.H drafted the manuscript, and all other authors contributed to it. The corresponding author is the guarantor and declares that all authors meet authorship criteria and that no other authors meeting the criteria have been omitted.

## Supplementary file 1. Search strategy

### Search strategy

(((cleft* AND (lip* OR palat* OR alveol*)) OR “alveolar cleft “) AND ((platelet* AND (fibrin* OR plasma* OR concentrat*)) OR PRF OR P-PRP OR L-PRP OR P-PRF OR L-PRF “) AND (((randomized controlled trial [pt] OR controlled clinical trial [pt] OR randomized [tiab] OR placebo [tiab] OR drug therapy [sh] OR randomly [tiab] OR trial [tiab] OR groups [tiab]) NOT (animals [mh] NOT humans [mh]))))

## Bibliography

1. Guo J, Li C, Zhang Q, et al. Secondary bone grafting for alveolar cleft in children with cleft lip or cleft lip and palate. Cochrane Database Syst Rev. 2011;2011(6). doi:10.1002/14651858.CD008050.pub2

2. Waite PD, Waite DE. Bone grafting for the alveolar cleft defect. Semin Orthod. 1996;2(3):192–196. doi:10.1016/S1073-8746(96)80014-4

3. Boyne PJ, Sands NR. Secondary bone grafting of residual alveolar and palatal clefts. J Oral Surg. 1972;30(2):87–92.

4. Wu C, Pan W, Feng C, et al. Grafting materials for alveolar cleft reconstruction: a systematic review and best-evidence synthesis. Int J Oral Maxillofac Surg. 2018;47(3):345–356. doi:10.1016/j.ijom.2017.08.003

5. Khojasteh A, Kheiri L, Motamedian SR, Nadjmi N. Regenerative medicine in the treatment of alveolar cleft defect: A systematic review of the literature. J Cranio-Maxillofacial Surg. 2015;43(8):1608–1613. doi:10.1016/j.jcms.2015.06.041

6. Marukawa E, Oshina H, Iino G, Morita K, Omura K. Reduction of bone resorption by the application of platelet-rich plasma (PRP) in bone grafting of the alveolar cleft. J Cranio-Maxillofacial Surg. 2011;39(4):278–283. doi:10.1016/j.jcms.2010.04.017

7. Oyama T, Nishimoto S, Tsugawa T, Shimizu F. Efficacy of Platelet-Rich Plasma in Alveolar Bone Grafting. J Oral Maxillofac Surg. 2004;62(5):555–558. doi:10.1016/j.joms.2003.08.023

8. Lee C, Nishihara K, Okawachi T, Iwashita Y, Majima HJ, Nakamura N. A quantitative radiological assessment of outcomes of autogenous bone graft combined with platelet-rich plasma in the alveolar cleft. Int J Oral Maxillofac Surg. 2009;38(2):117–125. doi:10.1016/j.ijom.2008.11.019

9. Luaces-Rey R, Arenaz-Búa J, López-Cedrún-Cembranos JL, et al. Is PRP useful in alveolar cleft reconstruction? Platelet-rich plasma in secondary alveoloplasty. Med Oral Patol Oral Cir Bucal. 2010;15(4):619–623. doi:10.4317/medoral.15.e619

10. Ouzzani M, Hammady H, Fedorowicz Z, Elmagarmid A. Rayyan—a web and mobile app for systematic reviews. Syst Rev. 2016;5(1):210. doi:10.1186/s13643-016-0384-4

11. Review Manager (RevMan). Published online 2014.

12. Moher D, Liberati A, Tetzlaff J, Altman DG. Preferred reporting items for systematic reviews and meta-analyses: the PRISMA statement. BMJ. 2009;339(jul21 1):b2535–b2535. doi:10.1136/bmj.b2535

13. Higgins JPT, Thomas J, Chandler J, et al., eds. Cochrane Handbook for Systematic Reviews of Interventions. Wiley; 2019. doi:10.1002/9781119536604

14. Sterne JAC, Savović J, Page MJ, et al. RoB 2: a revised tool for assessing risk of bias in randomised trials. BMJ. Published online August 28, 2019:4898. doi:10.1136/bmj.l4898

15. Group G working. GRADEpro Guidance Development. Published online 2015. tool.gradepro.org

